# Interpretable Radiomics Method for Predicting Human Papillomavirus Status in Oropharyngeal Cancer using Bayesian Networks

**DOI:** 10.1101/2022.06.29.22276890

**Authors:** Oya Altinok, Albert Guvenis

## Abstract

**Objectives:** To develop a simple interpretable Bayesian Network (BN) to classify HPV status in patients with oropharyngeal cancer.

**Methods:** Two hundred forty-six patients, 216 of whom were HPV positive, were used in this study. We extracted 851 radiomics markers from patients’ contrast-enhanced Computed Tomography (CT) images. Mens eX Machina (MXM) approach selected two most relevant predictors: sphericity and max2DDiameterRow. The area under the curve (AUC) demonstrated BN model performance in 30% of the data reserved for testing. A Support Vector Machine (SVM) based method was also implemented for comparison purposes.

**Results:** The Mens eX Machina (MXM) approach selected two most relevant predictors: sphericity and max2DDiameterRow. Areas under the Curves (AUC) were found 0.78 and 0.72 on the training and test data, respectively. When using support vector machine (SVM) and 25 features, the AUC was found 0.83 on the test data.

**Conclusions:** The straightforward structure and power of interpretability of our BN model will help clinicians make treatment decisions and enable the non-invasive detection of HPV status from contrast-enhanced CT images. Higher accuracy can be obtained using more complex structures at the expense of lower interpretability.

**Advances in Knowledge:** Determination of HPV status can be done by invasive laboratory techniques, which poses a potential risk to patients. Radiomics-based methods are non-invasive but are usually difficult to use because they are generally not interpretable. Therefore, there is a growing need to develop a non-invasive radiomics method that is simple and interpretable. This work accomplishes this objective while pointing out the limitations.

## I. INTRODUCTION

The availability of big open-source data in medical imaging allows for developing new approaches in personalized treatment for cancer. Advanced machine learning techniques for therapy selection and risk assessments have been increasingly appreciated in the era of precision oncology^1^. Support vector machines^2^, logistic regression^3^, random forest^4^ are some examples of the most studied techniques in this context. However, many of these techniques are not clearly understood by physicians at the stage where they are trying to rely on machine-based predictions. The explanatory power and simplicity of the model are an urgent need in the healthcare system to persuade health care providers to use predictive machine learning methods. Therefore, predictive modelling with Bayesian networks (BN) has advantages over regression-based models ^5^.

This study investigated an interpretable and straightforward BN model for predicting human papillomavirus (HPV) in oropharyngeal cancer. Head and Neck cancer (HNC) ranks 6^th^ among all cancer types ^6^. Oropharyngeal and oral cavity cancers, the subtype of head and neck cancers, affect 3% and 2% of men and women, respectively, in the USA^7^. Although there are more than 100 known subtypes of HPV, HPV type 16 (HPV-16) predominates in this specific type of cancer^8^. Furthermore, HPV-related cases in this specific subregion have more than doubled over the past four decades^9^, and, today up to 63% of oropharynx cancers are found to be HPV-infected^10^. Therefore, besides general risk factors such as smoking, alcohol, oral hygiene, diet, and genetics, HPV-16 infection has been recognized as the leading cause of cancer in this region^8^.

The presence of HPV infection can be identified by invasive laboratory techniques such as p16 immunohistochemistry (IHC) and PCR. According to American Joint Commission on Cancer (AJCC), p16 IHC was proposed as a prognostic test for oropharyngeal cancer staging at 8^th^ edition^11^. However, this may cause an extra burden on clinical applications and waste time. This shows us the urgent importance of identifying HPV status using non-invasive methods.

Radiomics biomarkers have been appreciated to identify HPV positivity in head and neck cancer^12,13^. HPV infected cancers have been reported radiologically different from non-infected tumors^13–15^. One group^16^ studied textural analysis of contrast-enhanced CT images of oropharyngeal cancer patients and showed statistically significant textural differences between HPV positive and negative tumors.

Determination of HPV-16 status completely changes the treatment to be applied to the patient. For example, numerous studies reported that in patients with oropharyngeal cancer who tested positive for HPV, the success rate of response to treatment increased. In fact, the predicted life expectancy compared to patients with HPV negative testing was found higher was found higher^7,10,17^.

Several machine-based inferential techniques^18–20^ attempted to determine the status of HPV in oropharyngeal cancer. However, many of them are far from clinical use because of the complexity of the methods and the lack of explanatory power of the models.

Bayesian networks are directed acyclic graphs (DAG) that reveal probabilistic relationships between variables^21^. Nodes and arcs are two elements of DAG. Nodes represent radiomic biomarker variables while arcs connect nodes and show probabilistic associations between nodes. Regarding quantitative computations of the DAG, conditional probabilities are defined for each variable (node). In addition, DAG is a way of visual representation of the model. In terms of user-model interaction, visual representation is a straightforward explanation of the model^22^. Therefore, network analysis for predictive modeling provides interpretability to professionals seeking to understand the interrelationships of variables.

There are three ways to identify the structure of DAG. One way is expert knowledge, the second way is to learn from the data, and the third way is the combination of the first two. However, it is more challenging to learn DAG from data, as the complexity of computations and possible structures (DAGs) increases as the number of variables increases^23^.

Here, we developed a straightforward BN structure to determine HPV positivity to guide medical practitioners in designing personalized therapy for patients with oropharyngeal cancer.

## II. MATERIALS AND METHODS

Figure 2 represents the schematic overview of our methodology. Figure S1 (Supplementary materials) shows the overall workflow. A Support Vector Machine (SVM) based method was also implemented for comparison purposes (Figure S2).

### Data Set

The dataset used in this study consists of 246 patients. Patient selection diagram is shown in Figure 1. and patient characteristics are given in Table 1. diagnosed with oropharyngeal cancer, and contrast-enhanced Computed Tomography (CT) images (with segmentation labels) were retrieved from The Cancer Imaging Archive (TCIA)^24–26^. HPV labels are available for all patients, p16 protein-positive or negative, in the relevant data set. Gross primary tumor volume (GTVp) and gross nodal tumor volume (GTVn) are the regions of interest (ROI) segmented by experts manually, as described in ^24^. In ^27,28^ only GTVp is considered for radiomic extraction. Therefore, since our concern is to determine HPV status, we also consider ROIs defined as GTVp in this dataset.

**Table 1.** Patient characteristics of all included patients. Continuous variables are represented as means ± standard deviation.

| Characteristics | Statistics (N=246) |
| --- | --- |
| Age(mean) (yr) | 58.28 ± 9.259 |
| Sex (No.) (%) |  |
| Female | 44 (17.9%) |
| Male | 202 (82.1%) |
| Smoking (No.) (%) |  |
| Current | 60 (24.4%) |
| Former | 97 (39.4%) |
| Never | 89 (36.2%) |
| HPV Status (No.) (%) |  |
| Negative | 36 (14.6%) |
| Positive | 219 (85.4%) |
| T Category (No.) (%) |  |
| 1 | 63 (25.6%) |
| 2 | 97 (39.4%) |
| 3 | 50 (20.3%) |
| 4 | 36 (14.6%) |
| N Category (No.) (%) |  |
| 0 | 23 (9.3%) |
| 1 | 31 (12.6%) |
| 2 | 186 (75.6%) |
| 3 | 6 (2.4%) |

**Figure 1.**
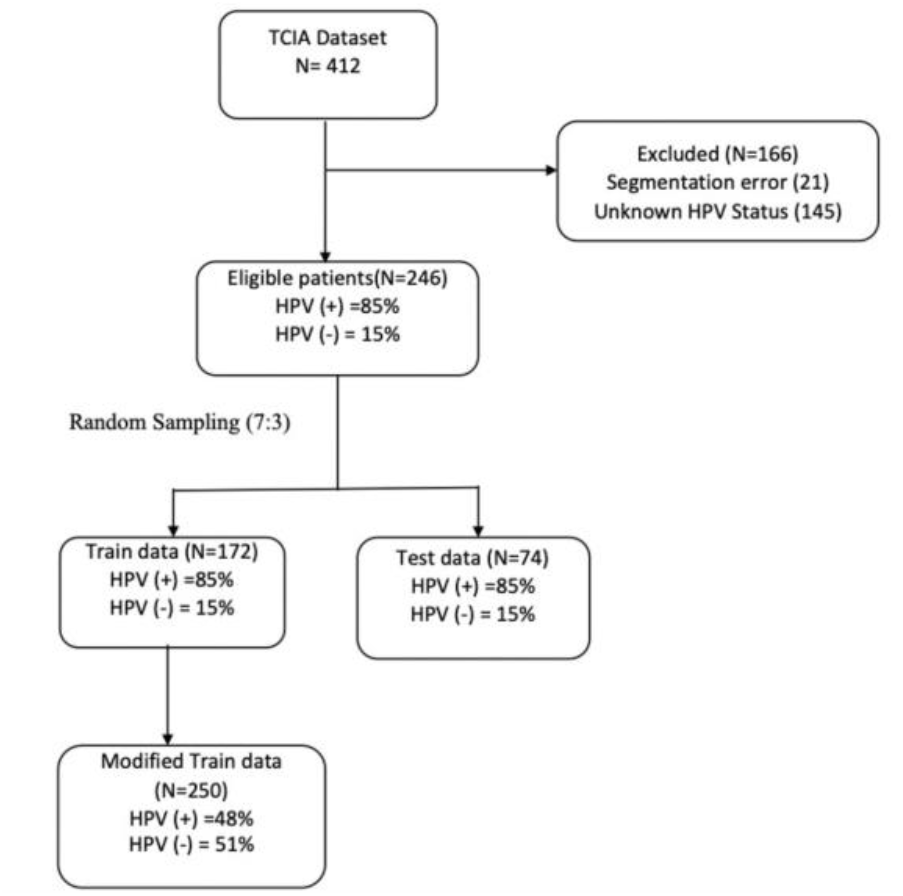
This figure gives the patient selection diagram based on the data given in ^24–26^.

### Image Preprocessing

Some subjects were not included because ROIs associated with the images were not found. All patient images were resampled to 1mm x 1mm x 1mm spatial resolution to avoid varying voxel sizes^24^ and make them comparable. The B-spline interpolation^29^ method was used to find new values. Resampling and interpolation were done in 3D Slicer software (version 4.11.2)^30^.

### Feature Extraction

Feature extraction is converting medical images (ROI) into quantitative features. After image preprocessing, radiomics features were extracted by using 3D Slicer software (version 4.11.2)^30^ with PyRadiomics^31^ extension. Grey-level discretization was performed by setting the bin width to 10 to reduce heterogeneity^32^ in the 3D slicer. The radiomic features of original images and wavelet transform images were extracted in 5 different categories: shape, intensity statistics, gray level co-occurrence matrix (GLCM), gray level run length matrix (GLRLM), and gray level size zone matrix (GLSZM). Full descriptions of these radiomic categories can be found in ^31^.

### Data Preprocessing

Z-score normalization was applied to high dimensional data with 851 radiomics features. Continuous data were discretized into four levels using interval method of the to improve classification performance^33^. The dataset was split for 70% training and 30% testing. However, there was inequality in the number of HPV cases. The proportion of patients who are HPV negative was approximately 15%, and the rest were positive in the dataset. Therefore, due to the inequality in HPV cases, Random Over-Sampling Examples (ROSE)^34^ in R^35^ was applied to the training dataset to generate synthetic data. The final training data consisted of 250 patients with approximately equal HPV positive and negative patient rates. The test data was not subject to ROSE.

### Feature Selection

We applied two statistical methods to obtain extracted data’s most informative radiomics features. First, an independent t-test was performed between HPV-positive and negative categories in SPSS 27, yielding the 25 most valuable features. The data were considered significant with a p-value of <0.05.

Then, we wanted to acquire the smallest subset of features with the highest predictive performance since we want structure of our model to be simple. As the number of variables in the model increases, the complexity of the model increases, and its intelligibility decreases^36^. Therefore, we used Mens eX Machina (MXM) package^37^. MXM is a feature selection package that can handle various types of target variables, and it is also successful in finding the minimal subsets of features for predictive models. To do that, we applied the Max-Min Parent-Child (MMPC) algorithm to identify the minimal subset of predictive features. MMPC is a Markov blanket filter-based algorithm that can identify parents and child of the target variable. Since Markov Blanket of the target variable has all necessary information for the target, additional variables are not needed. We set *test=testIndLogistic*, which is binary logistic regression for conditional independence test since our target variable is binary.

### Learning the Bayesian Network Structure and Parameters

We used R^35^ package *bnlearn*^38^ to learn BN structure with two predictors: Sphericity and Max2DDiameterRow. HPVStatus is a binary variable, which shows whether a patient has an HPV infection or not. We used the hill-climbing algorithm by means of a score-based structure learning approach, to learn the network structure. Then, parameters were learned using the *bn*.*fit* function. By doing so, conditional probabilities of nodes (variables) were obtained. Only modified training data was used to learn the structure and the parameters.

### Cross-validation

We performed 5-fold cross-validation on the training dataset to evaluate the performance of the predictive model. Training data was partitioned into five subsets with equal sizes. The prediction error for HPVStatus was set as a loss function. The resulting structure consists of the *test, fitted*, and *learning* elements. First, we predicted the values for test cases in each fold k. Next, the *fitted* element was converted to a gRain object, and then the k-th test subset was created.

### Performance Measurement of the Predictive Model

#### Training data

The area under the curve (AUC) evaluated model performance. We used the *caTools* package (Version: 1.18.2) to evaluate our model’s performance.

#### Test data

We used the *gRain*^39^ package to perform posterior inferences of the fitted BN. First, BN parameters were learned (on training data). Then, we used *test data* to predict HPV class. The area under the curve (AUC) evaluated model performance.

### Measurement of Arc Strength

We measured the relationship of the connected nodes using the *arc*.*strength* function, and *criterion = X2* was set. The strength represents a probabilistic relationship of an arc of BN. It computes strength for each arc and keeps fixed the rest of the network structure. The criterion is a conditional independence test, and the strength measures a p-value. Therefore, more robust relationships are lower p-values and versa. Furthermore, we computed the strength of every possible arc. This function computes both frequencies of arcs presented after bootstrap samples and the direction of arcs.

### Statistical Analysis

Statistical analyses were done using IBM SPSS 27. Independent samples t-test analysis was performed. All tests were two-tailed.

## III. RESULTS

Two hundred forty-six patients with oropharyngeal cancer were included in this study. Two hundred sixteen patients were HPV positive, and the rest were negative. The patients were split %70 and %30 as training (172 patients) and testing (74 patients). Modified training data set included 115 HPV-positive patients and 135 HPV-negative patients.

First, by applying an independent sample t-test, 25 radiomic features were selected between HPV-positive and HPV-negative cases. Then, two radiomics features were selected by Mens eX Machina (MXM), a feature selection method in R. Sphericity and Max2DDiameterRow were identified after the feature selection process. These two radiomic biomarkers are shape features of the tumor. Figure 3A shows that HPV-positive tumors were more spherical than HPV-negative tumors, and Max2DDiameterRow (Fig 3B) was more significant in HPV-negative tumors. Our hypothesis was that these identified radiomic features, extracted from CT images, can predict HPV status noninvasively in oropharyngeal cancer patients.

**Figure 2.**
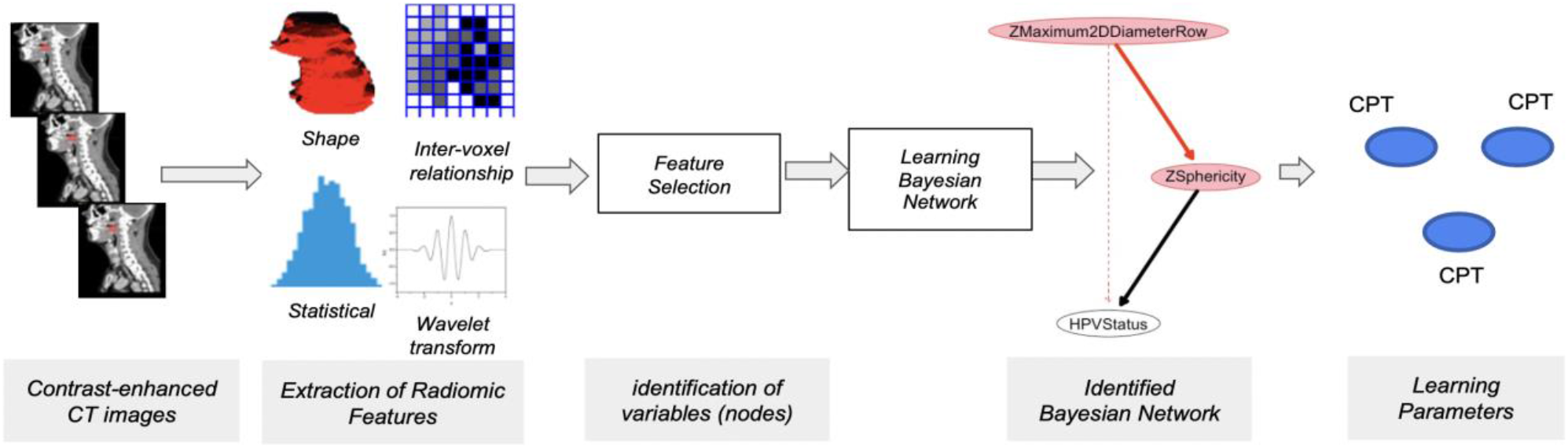
Schematic view of the methodology. CPT depicts conditional probabilities between nodes.

**Figure 3.**
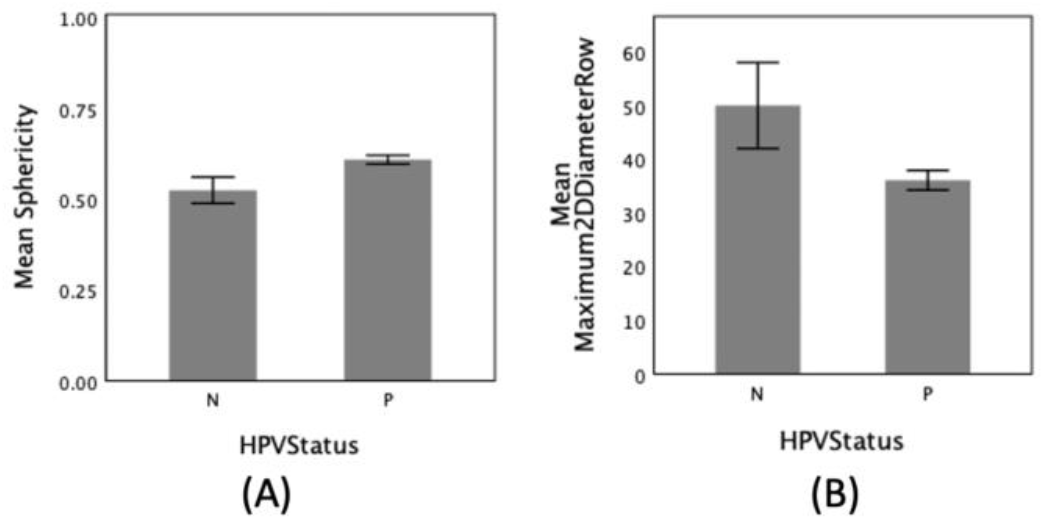
shows statistical significance between HPV positive and negative patients. P represents positive cases and N represents negative cases. (A) Comparison of mean sphericity (p<0.001), (B) Comparision of mean Max2DDiameterRow (p<0.001).

Therefore, we developed a straightforward BN structure to determine HPV positivity (Fig. 4). In the BN structure, directed arcs show the casual relationship of nodes. Red lines represent the negative association between nodes, while the black line represents the positive association. The thickness of an arc represents the strength of a connection between nodes.

**Figure 4.**
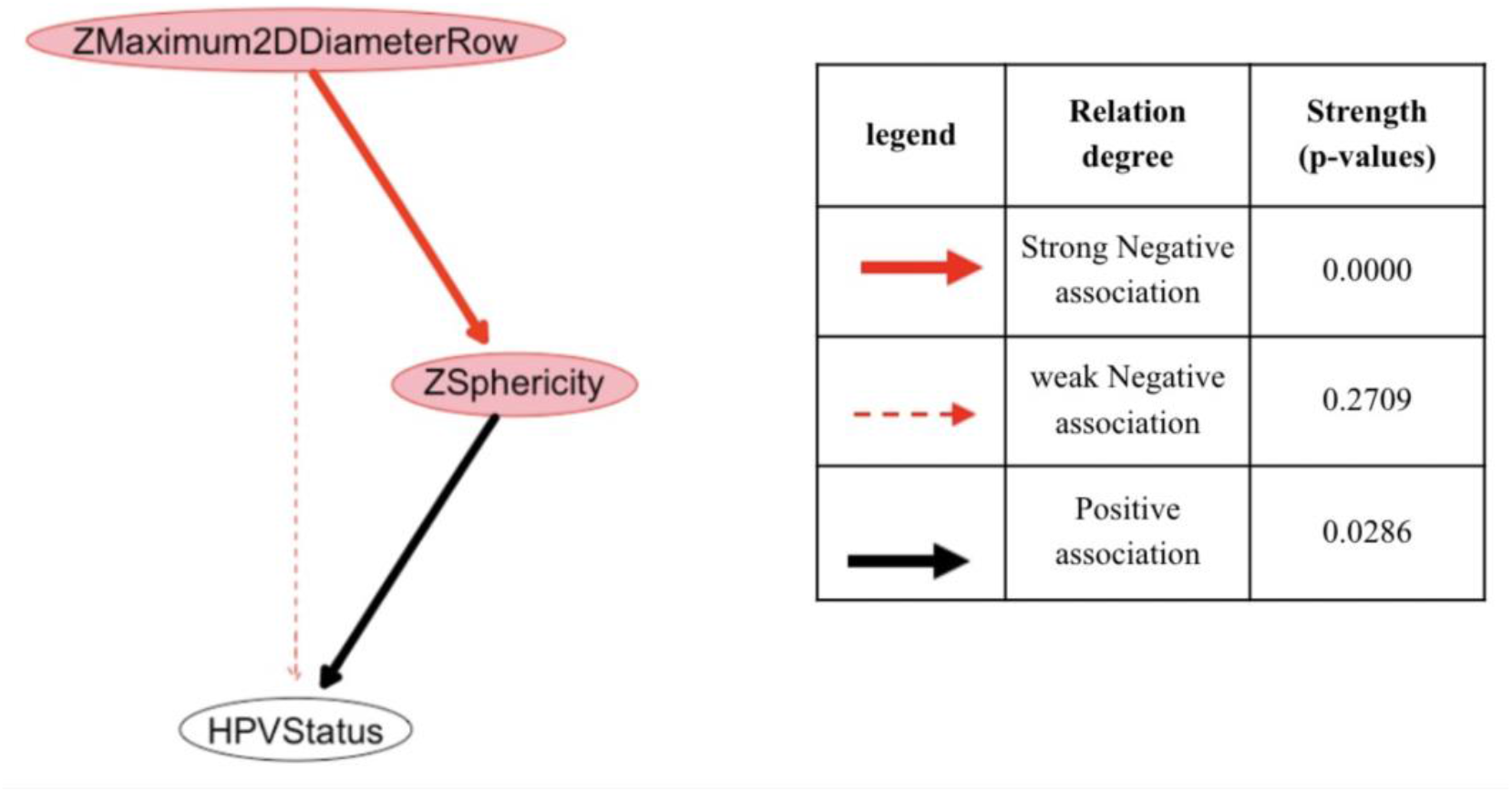
Predictive BN model structure. Pink nodes represent predictor values, and HPVStatus node is a class variable that classifies HPV status. The directed arcs show the probabilistic dependencies between nodes. Strength measures a p-value of an arc that expresses probabilistic relationships.

### Analyzing of the Network Structure and its parameters

Identified continuous variables (predictors) were discretized using the *interval* method into four levels. Modified training data set were used to learn the BN structure and parameters. We obtained our predictive BN structure from the training dataset using a hill-climbing algorithm. The multinomial log-likelihood (loglik) score calculated BN. Sphericity and Max2DDiameterRow are called the parents node of HPVStatus in the network. These parent nodes of the structure contain predictive parameters of the HPVStatus node.

We analyzed the arc strengths of the network (Fig. 4) to evaluate the probabilistic relationship of nodes (variables). The arc from Max2DDiameterRow to Sphericity shows a robust negative association (p-value < 0.0001), while the arc from Max2DDiameterRow to HPVStatus has a weak association (p-value = 0.27). On the other hand, arc strength from Sphericity to HPV status is strong with p-value = 0.028.

After the BN structure was identified, the parameters of the network were learned. First, we fit parameters of the network using their bayes parameter estimation. Thus, the conditional probability tables for nodes and the class probabilities were determined (Table 2). This allows us to predict HPV probabilities for new observations.

**Table 2.** The table of conditional probabilities *P* (HVPStatus | Sphericity, Max2DDiameterRow) The data were discretized into four levels. Sphericity (0.596 ± 0.967) and Max2DDiameterRow (38.23 ± 16.31), here, are actual values for illustration purposes. They are represented as means ± standard deviation.

| Sphericity = [0.289898,0.41956] |  |  |  |  |
| --- | --- | --- | --- | --- |
| Maximum2DDiameterRow | [12.1655,37.0403] | (37.0403,61.9151] | (61.9151,86.7899] | (86.7899,111.665] |
| HPV(Positive) | 0.99689441 | 0.001039501 | 0.001557632 | 0.001039501 |
| HPV(Negative) | 0.00310559 | 0.998960499 | 0.998442368 | 0.998960499 |
| Sphericity = (0.41956,0.549222] |  |  |  |  |
| Maximum2DDiameterRow | [12.1655,37.0403] | (37.0403,61.9151] | (61.9151,86.7899] | (86.7899,111.665] |
| HPV(Positive) | 0.3333623 | 0.3555756 | 0.2353914 | 0.0006242197 |
| HPV(Negative) | 0.6666377 | 0.6444244 | 0.7646086 | 0.9993757803 |
| Sphericity = (0.549222,0.678884] |  |  |  |  |
| Maximum2DDiameterRow | [12.1655,37.0403] | (37.0403,61.9151] | (61.9151,86.7899] | (86.7899,111.665] |
| HPV(Positive) | 0.7758323 | 0.6944107 | 0.998442368 | 0.5 |
| HPV(Negative) | 0.2241677 | 0.3055893 | 0.001557632 | 0.5 |
| Sphericity = (0.678884,0.808546] |  |  |  |  |
| Maximum2DDiameterRow | [12.1655,37.0403] | (37.0403,61.9151] | (61.9151,86.7899] | (86.7899,111.665] |
| HPV(Positive) | 0.757527 | 0.444483 | 0.5 | 0.5 |
| HPV(Negative) | 0.242473 | 0.555517 | 0.5 | 0.5 |

### Interpretability and Explainability of the Model

Class variable (HPV status) can be determined with maximum probability from the BN with joint probability distribution (JPD). JPD allows us to do probabilistic reasoning to estimate the value of a particular node given the values of other nodes. Sphericity and Max2DiameterRow are two input nodes of our BN. To estimate a probability of outcome, we can set evidence for both nodes and query the probability of HPV being positive or negative. Table 2 shows the conditional probabilities table of our model that is learned from the modified training data. For example, if sphericity = [0.289898,0.41956] and Max2DDiameterRow = [12.1655,37.0403], it is highly probably that a patient is HPV positive (99.68%) (Table 2). In addition, the visual graph of the BN model (Figure 4) can be considered the model’s explanatory power. As explained in the previous result section (analyzing of the network structure), our simple BN model can visually explain the interrelationship of radiomic features.

### The Performance of the BN model on the training dataset

We computed the performance of our predictive BN model on the modified training dataset based on cross validation (methods). On the training dataset, the AUC of predictions is 0.73-0.90, with an average AUC of 0.78. ROC curve of BN with training dataset is shown in Figure 5(A).

**Figure 5.**
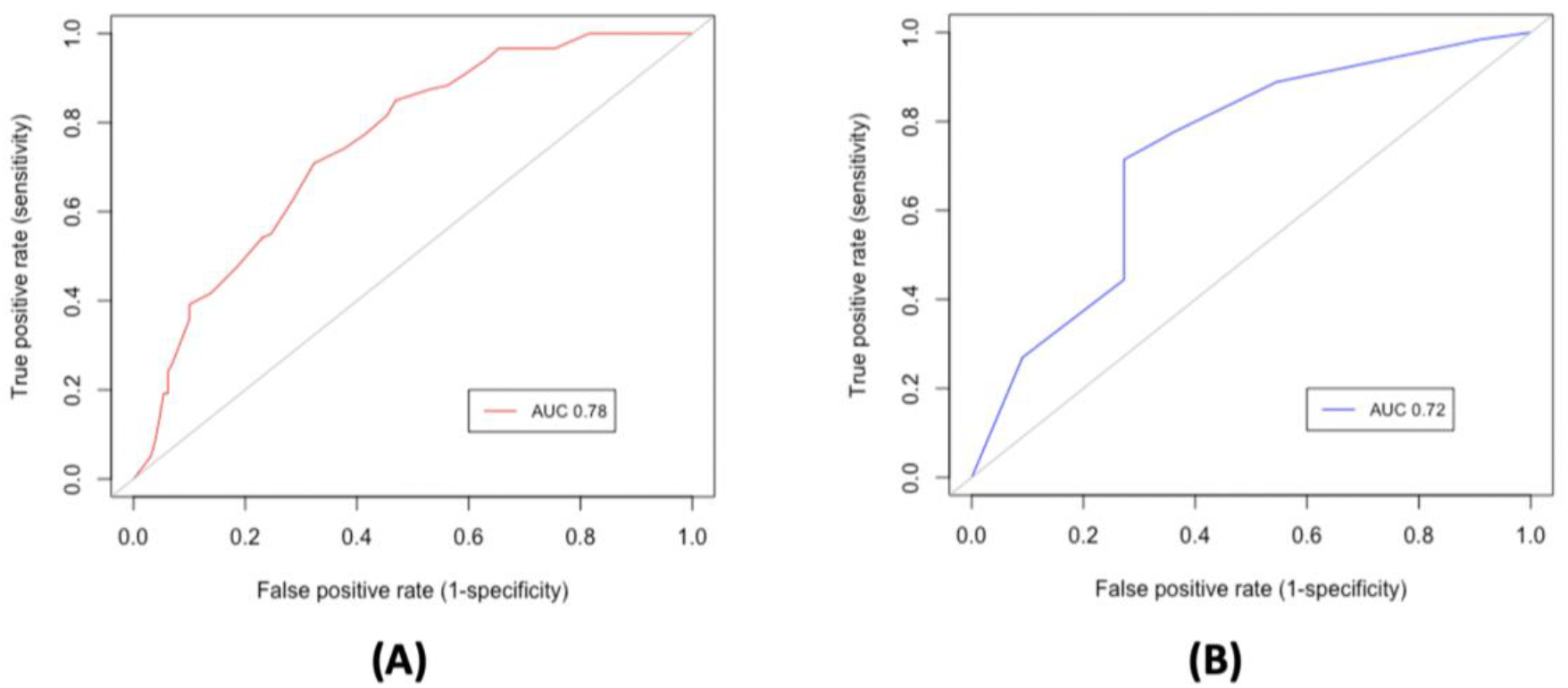
(A) ROC curve of BN as shown in Fig.3 based on cross validation with the modified training dataset. (B) ROC curve of BN with the test dataset.

### The Performance of the BN model on the test dataset

To evaluate our BN model, we measured its performance on the test dataset. Trained BN was used in this context. We did not change any parameters of trained BN, and testing data had no contribution to structure learning and parameter learning of the BN. The model performance tested on test dataset had an AUC of 0.72. ROC curve of BN with the testing dataset is illustrated in Figure 5 (B).

## IV. DISCUSSION

Understanding the degree of relationships between variables (nodes) is a crucial advantage over frequency-based machine learning methods. However, for many methods it is hard to understand how a model handles classification. Artificial neural networks (ANNs) and support vector machines (SVMs) are two well-known examples of black-box classifiers ^40^. In addition, logistic regression is also considered a black box in a very recent study ^41^. Cynthia Rudin, in her article ^42^, warns about the use of “black-box machine learning models” for risky decisions and emphasizes the need for a shift to “interpretable models”. Mihaljević et al. ^36^ suggest that BN is a suitable approach for interpretable machine learning modelling.

In terms of interpretability of a model, transparency is used as a counter term to the black box^43^. Barredo Arrieta et al. ^44^ evaluates different machine learning methods under the concept of transparency. Linear and logistic regression models are considered transparent, but they require post-hoc analyses to make them interpretable, and their interpretability may depend on who will be the user ^44^. However, classifiers should be easy to understand, as they are intended to assist professionals in diagnosis or prognosis. Table 3 compared several machine learning models for HPV status prediction in CT of head and neck cancer patients. Although many of these result in better performance (AUC) than our model, the problem of interpretability arises as suggested in ^41,44^.

**Table 3:** Comparison of the proposed method with machine learning models for HPV status prediction in CT of head and neck cancer patients. N represents the total number of features used in the model.

|  | Number of patients | Classifier | Performance (AUC) |  | Features |
| --- | --- | --- | --- | --- | --- |
|  |  |  | Train | Test |  |
| Yu et al.<sup>20</sup> | 315 oropharynx cancer patients | Logistic regression | 0.753 | 0.915 | (N=2 features) Shape Features: MeanBreadth & SphericalDisproportion |
| Bagher-Ebadian et al.<sup>45</sup> | 187 oropharyngeal cancer patients | General Linear Model | 0.849 | 0.869 | (N=12 features) GLCM – Contrast, Energy, Entrophy<br>Shape Features -MeanBreadth & SphericalDisproportion<br>DOST- Energy1,2,3,4,6,7,8,9 |
| Bogowicz et al.<sup>28</sup> | 149 HNSCC<br>-Oropharynx (100)<br>-Other sites (49) | Multivariable logistic regression | 0.85 | 0.78 | (N=4 features) LLL standard deviation, LLL small zone high gray-level emphasis, HHL difference entropy, Coefficient of variation |
| Our work | 246 oropharynx cancer patients | Support Vector Machine (SVM) |  | 0.83 | (N=25 features) |
| Our work | 246 oropharynx cancer patients | Bayesian Network (BN) | 0.78 | 0.72 | (N=2 features)<br>Sphericity, Max2DDiameterRow |

This dataset was used for Medical Image Computing and Computer Assisted Intervention (MICCAI) grand challenge in 2016. The winner of the competition published their article^20^, in which they showed that their model (Table 3) performance resulted in a higher AUC (0.915) on test data, compared to the AUC of the training data (0.753). In addition to the issue of the model’s interpretability, this raises a question about their model stability. However, our model, together with its AUC (0.78 and 0.72, in training and test data, respectively) and interpretability capacity (see Table 2), demonstrates a robust and stable model for predicting HPV status.

Apart from measuring the model’s performance (AUC), BN has a strong interpretation (probabilistic reasoning) capacity of the model. For example, providing one or more evidence to the model allows the probability of having HPV to be calculated (Table 2). In the clinical aspect, this could allow medical practitioners to easily understand the results of the model and design personalized therapies.

Regarding to explainability, our BN can predict HPV status and reveal probabilistic associations between variables (nodes). For example, arrows entering node “HPVStatus” in Figure 4 explain the probability of HPV positivity by the extracted CT based shape characteristics. We identified two prominent variables: sphericity and Max2DDiametersRow. Sphericity ranges from zero to one and calculates the volume of the tumor compared to a sphere^46^. The closer the tumor volume is to zero, the farther from spherical and closer to one, the more spherical it is. Sphericity has been demonstrated in the study^47^ as a promising radiomic biomarker extracted from MRI images of breast cancer patients to predict treatment outcomes. Max2DDiameterRow, on the other hand, is defined as the largest binary Euclidean distance on the tumor surface^31^. The largest Max2DDiameterRow implies the complexity of tumor shape since tumor complexity has been shown to be associated with radiomic shape features^48^. One group ^49^ studied differences in imaging characteristics and showed that HPV-related tumors have well-defined borders in oropharyngeal cancers while HPV-negative tumors have ill-defined borders. In this context, this explains why HPV-positive tumors are more spherical and less complex.

HPV-positive oropharyngeal tumors are biologically^50^ and radiologically^14^ different and more manageable^51^ compared to non-positive patients with high overall survival rates^10^. Maniakas et. al ^52^ surveyed North America and found that 67% of patients had undergone HPV laboratory testing; however, only 58.3% of these tests were utilized to treat head and neck cancer patients. Therefore, it is necessary that HPV-positive patients be identified first to apply the appropriate treatment. While invasive techniques are common ways to test, expenses and time limits are the two main constraints ^52^ on why HPV testing is not routinely performed in clinics. Here, our CT-based radiomic model can provide physicians with further guidance on the need for external HPV screening. In addition, the BN model can give a probability of being HPV positive for a new patient image. For instance, in table 2, we calculated probabilities of a scenario that a patient has Max2DDiameterRow= (61.9151,86.7899] (3^rd^ level interval in table 2) and Sphericity = [0.289898,0.41956] (smallest interval), then the patient would have 0.15% of being HPV-positive. On the other hand, as tumor becomes more spherical, for instance (0.549222,0.678884] (3^rd^ level interval), with same max2DDiameterRow level, likelihood of being HPV-positive increases (99.8%).

### Limitations and Future Directions

We used a minimal subset of variables to learn the structure of our BN model. Since BN is based on Bayesian statistics, it can be easily updated with new evidence or expert knowledge (new features). Therefore, our model can be expanded with expert knowledge to identify clinical predictors that can improve our model reliability. In addition, further investigation can be done on a larger and independent external validation set to evaluate the predictive power of our BN model. Applying our model to larger datasets with more class homogeneity will be considered as future work since there is no optimal rule to solve the imbalance problem^53^.

The future use of our developed model as software or web-based tool in clinics can be a beneficial guide in deciding on treatments. Furthermore, our methodology may be used and evaluated for other types of cancer.

## V. CONCLUSION

Our study shows that the Bayesian network can predict HPV status from radiomic markers extracted from contrast-enhanced CT images. Bayesian network analysis is a useful predictive tool for HPV status. Additionally, the model can demonstrate probabilistic relationships between predictors (nodes). Unlike “black-box machine learning models”, BN is a transparent model which allows interpretability.

## Data Availability

All data produced in the present study are available upon reasonable request to the authors.

## Conflict of interest

Authors declare no conflict of interests.

## Notes

### Competing Interest Statement

The authors have declared no competing interest.

### Funding Statement

This study did not receive any funding.

### Author Declarations

RESTRICTED LICENSE AGREEMENT of THE CANCER IMAGING ARCHIVE gave approval for this work. Here is the link of aforementioned data set: https://wiki.cancerimagingarchive.net/display/DOI/Radiomics+outcome+prediction+in+Oropharyngeal+cancer#33948240036220c66a5a436f90e4a0b54367bfae

