## Supplementary material for "Interpretable Radiomics Method for Predicting Human Papillomavirus Status in Oropharyngeal Cancer using Bayesian Networks": Figure S1

%70

%30

Data Set

Performance of the model on modified train set

(AUC)

Rose() function to generate syntactic data N=250

**Structure Learning:**

hc() function to learn Bayesian network structure

**Figure S1.** shows the overall workflow. In this figure, the dashed lines represent the function applied to the dataset or the structure.

**Cross validation :**

bn.cv() function k=5,İss=0.1Method=bayes.

In this step bn.cv() also fits the parameters using “bayes” methos. Thus, CPT is also created in this step

**Parameter Learning:** bn.fit() function,uisng “bayes” method, iss=0.1)

Performance of the test set

(AUC)

Test Set

Learned BN Structure

Modified Data

Learned Parameters (CPT)

Train Set
