## Supplementary material for "Interpretable Radiomics Method for Predicting Human Papillomavirus Status in Oropharyngeal Cancer using Bayesian Networks": Figure S2

**I.MATERIALS AND METHODS**

**Support Vector Machine (SVM)**

A Support Vector Machine (SVM) based method was also implemented for comparison purposes. Student t-test was used to select the best 25 features.

**II. RESULTS**

**Selected features:** Flatness, MajorAxisLength, Maximum2DDiameterColumn, Maximum2DDiameterRow, Maximum3DDiameter, Sphericity, Maximum, Minimum, Range, Autocorrelation, JointAverage, SumAverage, DependenceEntropy, HighGrayLevelEmphasis, HighGrayLevelRunEmphasis, Maximum_A, Minimum_D, Range_D, JointAverage_D, SumAverage_D, DependenceEntropy_D, Minimum_H, Range_H, JointAverage_H, SumAverage_H

When using SVM and 25 features, the AUC was found 0.83 on the test data.

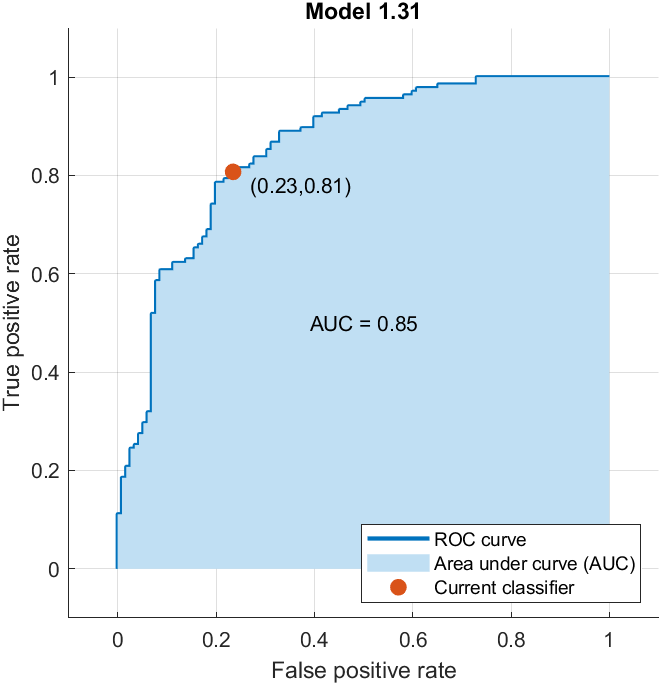

AUC =0.83

**Figure S2.** ROC curve of SVM model on the test dataset.
